# Autistic Neural ‘Shiftability’: The Distinct Pharmacological Landscape of the Autistic Brain

**DOI:** 10.1101/2024.11.08.24316969

**Authors:** Tobias P. Whelan, Lucas G. S. França, Mihail Dimitrov, Charlotte M. Pretzsch, Hester Velthuis, Andreia C. Pereira, Claire L. Ellis, Glynis Ivin, James L. Findon, Robert H. Wichers, Francesca M. Ponteduro, Johanna Kangas, Naoise Mulcrone, Nichol M. L. Wong, Dafnis Batalle, Nicolaas A. J. Puts, Eileen Daly, Declan G.M. Murphy, Gráinne M. McAlonan

## Abstract

Functionally inter-connected large-scale brain networks regulated by multiple neurotransmitter systems underpin complex human behaviour. A wide range of alterations in this neurofunctional landscape have been associated with autism. However, this evidence is mostly derived from cross-sectional analyses and thus assumes that any differences are ‘static’. Indeed, large sample sizes are required to identify reproducible baseline differences in functional connectivity between autistic and non-autistic individuals. Instead, we propose that what is different in autism is not necessarily the baseline functional connectivity of resting-state networks, but rather their responsivity or regulation by neurotransmitter systems. We tested this hypothesis using a uniform analytical framework to capture the modulation of functional connectivity by single doses of pharmacological probes targeting three major neurotransmitter systems implicated in autism, namely gamma-amino-butyric-acid (GABA), serotonin (5HT) and μ-opioid systems. Every drug challenge altered resting-state network connectivity differentially in the autistic compared to the non-autistic group. However, regardless of the neurotransmitter system probed, drug challenge elicited increases in between-network connectivity in autistic participants but minimal or decreased between-network connectivity in their non-autistic peers. There was no group difference in the responsivity of within-network connectivity. Thus, there is altered *responsivity* of neurotransmitter systems in the autistic brain. This has important implications for pharmacotherapy in autism because these neurotransmitter systems are the targets of several medications commonly prescribed to manage mental health conditions that frequently co-occur in autism. Investigating if these drug-induced ‘shifts’ in functional connectivity can help provide better targeted clinical interventions, will be important next translational steps.

**One Sentence Summary:** The autistic brain is differentially responsive to pharmacological modulation of the GABA, serotonin and μ-opioid neurotransmitter systems.

## Introduction

Autism is the result of biological differences in the brain. It is, however, complex and commonly complicated by the co-occurrence of mental health challenges. To date, no biological measure reliably separates autistic from non-autistic people^1^. This has been explained as a consequence of i) too broad a diagnostic definition of autism^2^ and/or ii) cohorts that are inherently heterogeneous; and/or iii) differences masked by mixing ages and genders^3^ and/or iv) a lack of harmonized methods^4^. Yet, perhaps more importantly, the brain is a dynamic organ and doesn’t simply function ‘at rest’; rather it *responds* to the modulatory effect of a *variety* of neurotransmitter systems. This is rarely examined in humans. Instead, most studies examine the brain at rest and not in response to the challenge of key neurotransmitter pathways. In our recent studies, we have begun to examine neural responsivity of the human brain to pharmacological challenge by targeting a specific neurotransmitter system, and found that autistic individuals respond differently than non-autistic individuals on selected neuroimaging parameters, which have included a ‘seed-based’ approach or during task conditions)^5^. However, we have not examined non-task dependent or ‘resting-state’ brain responsivity *across more than one neurotransmitter system (i.e. a variety of challenges)* using a uniform analytical framework.

Here we focus on functionally inter-connected large-scale brain networks which are generally accepted to underpin complex human behaviour^6^. Their within- and between-network connectivity can be captured using functional MRI (fMRI) techniques during ‘resting-state’, a condition uncomplicated by task-based demands. This systems-level communication arises through neuronal activity across several organisational scales. At the circuit-level, binding of receptors alters neuronal firing rate and modifies the ‘blood oxygen level dependent’ (BOLD) fMRI signal. Various alterations in functional connectivity have been associated with autism. However, identifying reproducible baseline differences in autism, for example incorporating the default mode and somatomotor networks^7^, has required extremely large sample sizes (e.g. in excess of n = 1800^7^), often pooling across multiple studies and cohorts for meta-analyses. Even then, these connectivity differences are only moderately associated with clinical traits of autism^8^. Moreover, to achieve such large samples, participants are often combined across age groups and baseline differences are especially limited when analyses are limited to autistic adults^3^. This demand for large sample sizes to identify only modest differences in baseline functional connectivity differences in autistic relative to non-autistic people, is also impractical for translating findings to clinical settings.

Cross-sectional designs also limit investigation of how neurotransmitter systems modulate functional connectivity in the human brain and prior studies have relied upon indirect correlational approaches to link measures of brain chemistry (for example, using PET or MR spectroscopy) to function. To provide direct experimental evidence for altered responsivity of functional networks in autism, we need to *change* the target system and observe *shifts* in connectivity which are distinct from non-autistic individuals. This concept underpins a series of studies adopting our ‘shiftability’ paradigm^5^, which involves pharmacologically challenging specific neurotransmitter systems and measuring the resultant shift in brain function across multiple organisational levels.

In our prior ‘shiftability’ studies of autistic and non-autistic adults we applied this paradigm to investigate several neurotransmitter systems with a range of compounds: acting at GABA receptors A (targeted by positive allosteric modulator AZD7325)^9^ and B (targeted by agonist arbaclofen)^10^, serotonin (5HT; targeted serotonin re-uptake inhibitor citalopram)^11^ and μ-opioid receptor (targeted by agonist tianeptine)^12^ pathways. We chose these neurotransmitter systems as they have been implicated in autism by genetic, post-mortem and preclinical studies^13,14^ (yet direct evidence for a functional difference is limited – see above) and they are also the targets of medications commonly prescribed to autistic people for co-occurring psychiatric and neurological conditions. In this study, we used the fMRI data collected across these individual studies to set-up a uniform analytical framework to test the hypothesis that the pharmacological responsivity of the autistic and non-autistic human brain is different.

First, we assessed the baseline group differences in within- and between-network functional connectivity which, based on extant literature, we predicted to be minimal between autistic and non-autistic participants at this sample size (total non-autistic, n = 67 and autistic, n = 54 across all study samples included)^3^. Next, we investigated within- and between-network responsivity in autistic and non-autistic participants by examining each pharmacological probe under a uniform analytical framework. Finally, we pooled within- and between-network shifts following drug challenge to establish any generalised response profile in autistic and non-autistic people.

## Materials and Methods

### Experimental Design

All component case-control studies included here were conducted in accordance with the Declaration of Helsinki at the Institute of Psychiatry, Psychology and Neuroscience (IoPPN), King’s College London (KCL) in London, UK. Our studies did not address safety or clinical efficacy and the UK Medicines and Health Regulatory Authority (MHRA) confirmed that these studies used a drug as a probe of neurotransmitter systems and none constituted a clinical trial of an Investigational Medicinal Product (IMP) as defined by the EU Directive 2001/20/EC. Nevertheless, our protocols were registered on clinicaltrials.gov for transparency. Ethical approval had been received from the appropriate Institutional Ethics Board or National UK Health Research Authority in the UK.

Data were collated across three distinct studies conducted during different time periods **(Table 1)**. All studies had a comparable design in which resting-state data were acquired following placebo and a single acute oral dose of a pharmacological probe (at maximum plasma concentration, t_max_) on separate visit days. The administration order was randomized across participants in all studies and the researcher and participant were blind to the dose order.

**Table 1.**
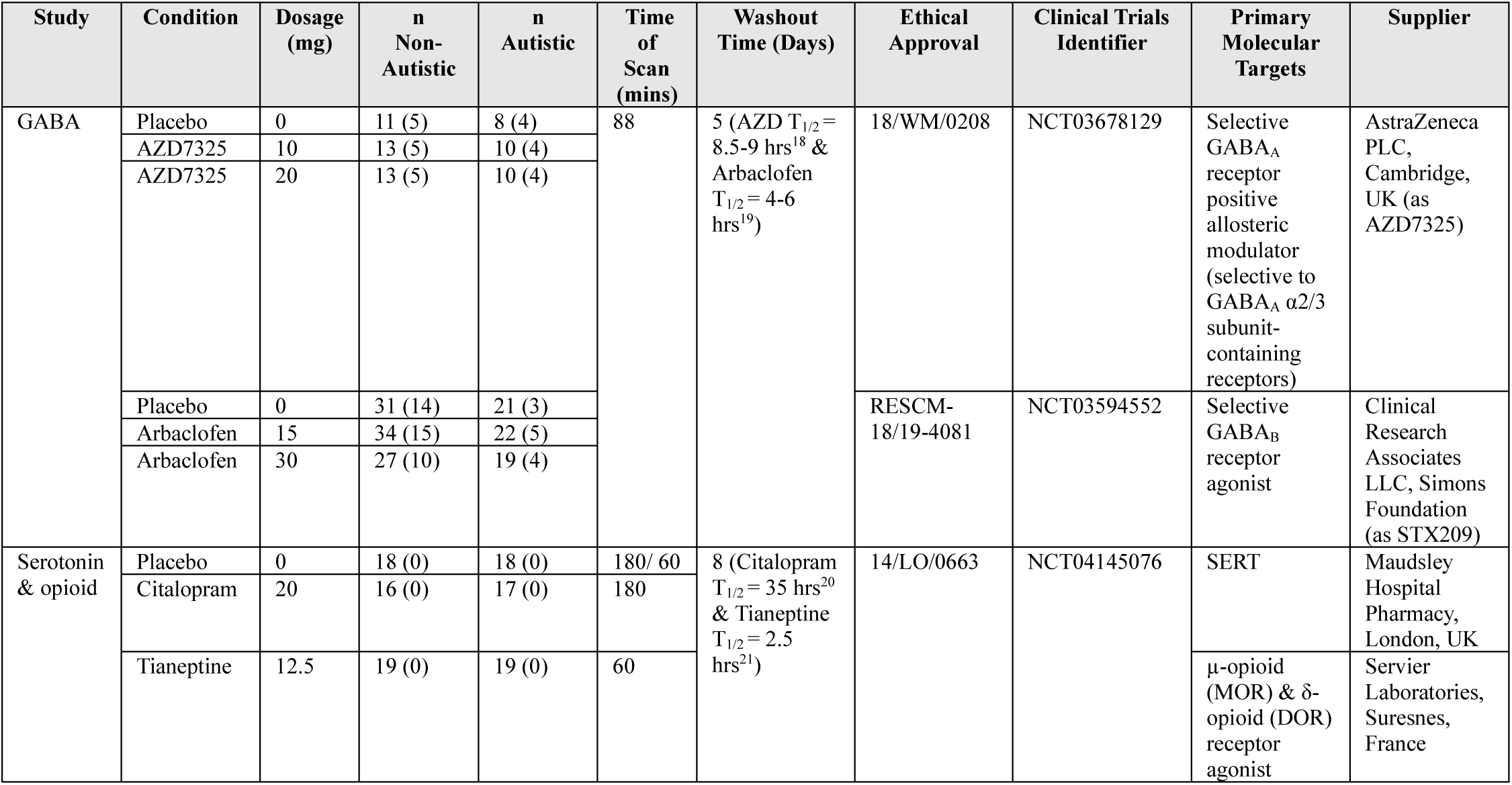
Studies included in the analyses. Values in parentheses indicate the number of female participants, note that only male participants were recruited for the serotonin and opioid study. ‘Time of scan’ refers to the approximate time of functional MRI scan post-administration of drug. ‘Washout time’ refers to the minimum time between repeat visits in which drug/placebo was administered and allowed sufficient time for drug to leave the system (half-life, T_1/2_). MRI data were acquired at maximum plasma concentration (T_max_). Given the different T_max_ of citalopram and tianeptine in the ‘serotonin & opioid’ study, placebo scans were acquired at the same time post-dose as either the citalopram (180 mins) or tianeptine condition (60 mins). In all studies, not all participants completed every study visit, this ‘dropout’ explains the discrepancy in the sample sizes between conditions (please see original reports for the details^15–17^).

### Participants

Participants recruited into the autism group had IQ>70 and a clinically confirmed Autism diagnosis as per the recruitment criteria of the parent study. The non-autistic participants were recruited to match the group age and IQ of the autistic participants. All participants gave written informed consent prior to being included in any experimental procedures. **Table 1** summarizes the parent studies providing data for these analyses. Further demographic information is included in **Supplementary Table 1**.

### MRI Data Acquisition

MRI acquisition information is provided in **Table 2** and includes the MR scanner, sequences to acquire both structural and functional scans, repetition time and echo time for each study included in analyses. Further information regarding MRI acquisition is in **Supplementary Table 2**.

**Table 2.** MRI acquisition information included in analyses. TR, repetition time; TE, echo time; ADNI GO, Alzheimer’s Disease Neuroimaging Initiative Grand Opportunities^22^ & echo planar imaging, EPI. Further information regarding MRI acquisition is in Supplementary Table 2.

| Study ID | MR Scanner | Scan | Sequence | TR (ms) | TE (ms) |
| --- | --- | --- | --- | --- | --- |
| GABA<sub>A/B</sub> | 3T General Electric Discovery MR750, Centre for Neuroimaging Sciences, King’s College London, UK | Structural | ADNI GO | 7312 | 3016 |
|  |  | Functional | EPI | 2500 | 12, 28, 44 & 60 |
| Serotonin & opioid |  | Structural | ADNI GO | 7312 | 3016 |
|  |  | Functional | EPI | 2300 | 12.7, 31 & 48 |

### MRI Data Preprocessing

For all studies the functional fMRI data was pre-processed using AFNI^23^ (AFNI v21.1.07 (https://afni.nimh.nih.gov/pub/dist/doc/htmldoc/#) and Anaconda v3.8 (https://anaconda.org/) and consisted of de-spiking; slice time correction; co-registration of fMRI to T1 structural scan; normalisation to standard (MNI152_T1_2009c) space; tissue segmentation; optimal combination of echoes and motion correction using multi-echo ICA (tedana v0.0.8)^24^; smoothing (6mm^3^); scaling (to percent signal change with µ = 0); band-pass filter (0.01 - 0.1Hz); regression of white matter and cerebrospinal fluid signal & censoring (if framewise displacement > 3mm, based on previous literature^8^ and/or if motion outlier (i.e. >2 σ)), additional information regarding head motion in the sample is in **Supplementary Table 3**. The raw and pre-processed MRI data was inspected and checked for quality issues using simple image visualisation in FSL’s FSLeyes in combination with examination of the reports that were automatically generated by AFNI. No scans were excluded from analyses for the AZD7325 (GABA_A_) study; 6 scans from non-autistic participants (two placebo, 15 mg and 30 mg sessions) and 6 scans from autistic participants (2 placebo, 15 mg and 30 mg sessions) were excluded for the arbaclofen (GABA_B_) study due to excessive signal loss; 3 scans from non-autistic participants (3 drug sessions) and 3 scans from autistic participants (3 drug sessions) were excluded from the serotonin (citalopram) study due to excessive motion and finally, 4 scans from non-autistic participants (2 placebo and 2 drug sessions) and 3 scans from autistic participants (2 placebo and 1 drug session) were excluded from the tianeptine (opioid) study due to excessive signal loss (i.e. motion artifacts).

### Functional Network Analysis

The pre-processed fMRI data for each drug was parcellated into 100 cortical brain regions of interest (ROIs) according to the Schaefer parcellation scheme^25^. The average time series in each parcel was used to construct functional connectivity matrices of Fisher-transformed Z values (or Fisher-transformed correlation coefficients) between every pair of ROIs for each individual scan using CONN v21a^26^. Each of the 100 ROIs were assigned into one of seven main functional networks according to the Yeo-7 atlas: default mode (DMN); frontoparietal (FPN); ventral attention (VAN) (or salience); dorsal attention (DAN); limbic (LN); somatomotor (SMN) & visual (VN) networks^25^. Based on this parcellation, within-network connectivity was defined as the mean connectivity between each ROI belonging to a Yeo-7 network. Between-network connectivity was defined as the mean connectivity between ROIs that comprises two different networks **(Fig. 1)**.

**Figure 1.**
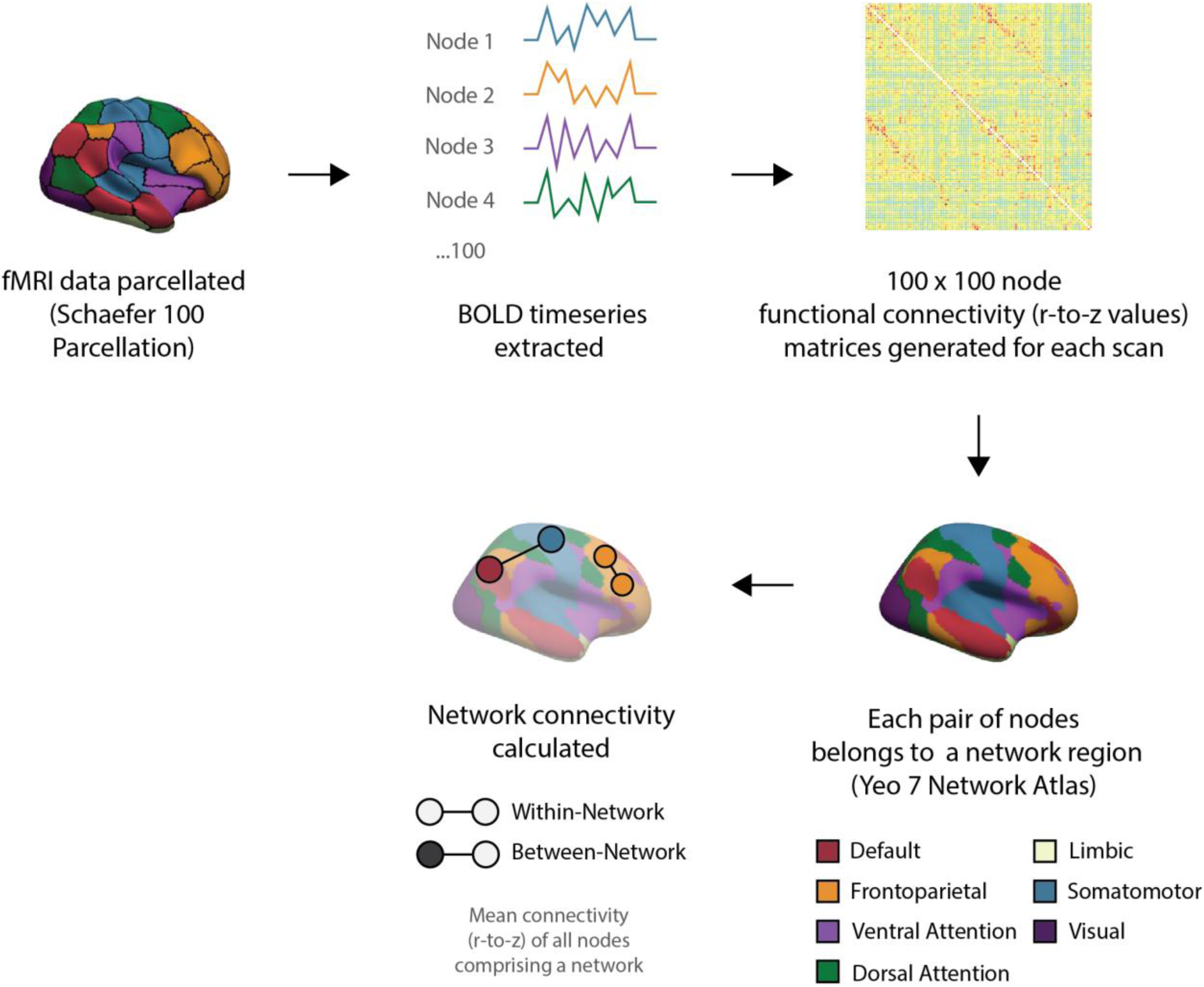
Schematic of the uniform resting-state network functional connectivity analysis pipeline applied to fMRI data from all studies. Data were parcellated into 100 cortical regions (REF); the average BOLD timeseries extracted from every node was then used to construct a functional connectivity matrix (of Fisher transformed Z values, r-to-z) between each of the 100 nodes for every individual scanning session. As each node is assigned a resting-state network (REF), within- and between-network connectivity can be calculated as the mean connectivity of the ROIs belonging to a given network or between ROIs comprising two different networks, respectively.

### Statistical Analyses

First, we tested the prediction that there would be no statistically significant differences within and between networks in the placebo condition between the autistic and non-autistic group. Then, for each drug we tested our main hypothesis that there would be a difference in the brain response to drug challenge in autistic and non-autistic individuals i.e. a dose × group interaction. To evaluate the statistical significance of the interaction between dose and group we fit a general linear mixed effects model (GLME) for all networks with two-sided permutation tests with 10,000 repetitions. The GLME used was given by the expression z̅ ∼ B0 + Bdose*group + (1|Subject ID) – with Subject ID accounting for the random effect. This approach accommodates missing data as not all participants had a complete dataset including placebo and all drug conditions. Significance was assessed with two-sided permutation tests featuring 10,000 GLME repetitions. We report p values obtained from the permutation tests uncorrected, highlighting those surviving multiple comparison correction using the False Discovery Rate method with α = 5%^27^. Dose × group interaction p values were corrected across all within- and between-networks for each pharmacological probe. While each study was conducted separately, a small number of autistic participants took part in two or more studies (n = 7, 5 male and 2 female) and were included in the analysis. We report the within-network results first, then the between-network results per pharmacological probe.

To investigate the extent to which any interactions were driven predominantly by a shift in the autistic group or non-autistic group, we estimated the Cohen’s d effect size^28^ for connectivity changes within- and between every functional network against placebo in each group separately; where our placebo baseline included every placebo fMRI scan from all studies (total non-autistic, n = 67 & autistic, n = 54). This is represented by equation XX, where z̅ represents the average Fisher’s Z for a given network and *σ*(Z) the standard deviation of such network:

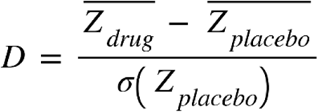

Finally, we examined whether drug challenge preferentially increases or decreases within- and between-network connectivity in the autistic brain, irrespective of pharmacological probe (and underlying neurotransmitter system targeted). To do this, we pooled the within- and between-network effect sizes generated in the analysis described above and then performed non-parametric Mann Whitney U tests to compare the aggregate within- and between-network connectivity shift between groups.

All statistical analyses were conducted in R version 4.1.1 (2021-08-10) (R Core Team 2021) with supporting packages ggplot2^29^, reshape2, dplyr, tidyr, tibble, shadowtext, lme4, and ptestr.

## Results

A comprehensive neuropharmacological profile of changes in the functional connectivity of cortical networks was produced by collecting resting-state fMRI data using four different pharmacological probes of the GABA_A_, GABA_B_, serotonin and μ-opioid systems. All fMRI data was parcellated into the same 100 cortical regions^25^. The resulting correlation matrices were Fisher-transformed (r-to-z values) and average connectivity calculated for all within- and between-network interactions using the previously defined atlas (each of the 100 cortical regions from the matrices belongs to a given Yeo resting-state network) of seven cortical functional networks: the default-mode (DMN), frontoparietal (FPN), ventral attention (VAN) (which is also referred to as the ‘salience network’), dorsal attention (DAN), limbic (LN), somatomotor (SMN) and visual networks (VN)^30^. An effect size (Cohen’s d) was then obtained for within and between each functional network to represent the magnitude of the ‘shift’ from baseline (i.e. inactive placebo).

### Autistic and non-autistic network connectivity is not significantly different at baseline

First, we examined the main effect of group to ascertain if there were baseline differences between autistic and non-autistic participants in our sample. Prior to correction for multiple comparisons (using the FDR method), only somatomotor-visual network between-network connectivity was significantly greater in the autistic group (p_unc_ = 0.007) (**Supplementary** Fig. 1). After correction for multiple comparisons, we observed no statistically significant differences between the groups (For the full main effect of group result at baseline (i.e. placebo) see **Supplementary Table 4**).

### Neurotransmitter systems differentially ‘shift’ network connectivity in the autistic brain

Below we report the within-network dose × group interactions (**Fig. 2**) and then the between-network connectivity dose × group interactions for each drug challenge (**Fig. 3**). Key findings of differential shifts in within- and between-network connectivity between the non-autistic and autistic group for each drug challenge are also summarised in **Table 3**.

**Figure 2.**
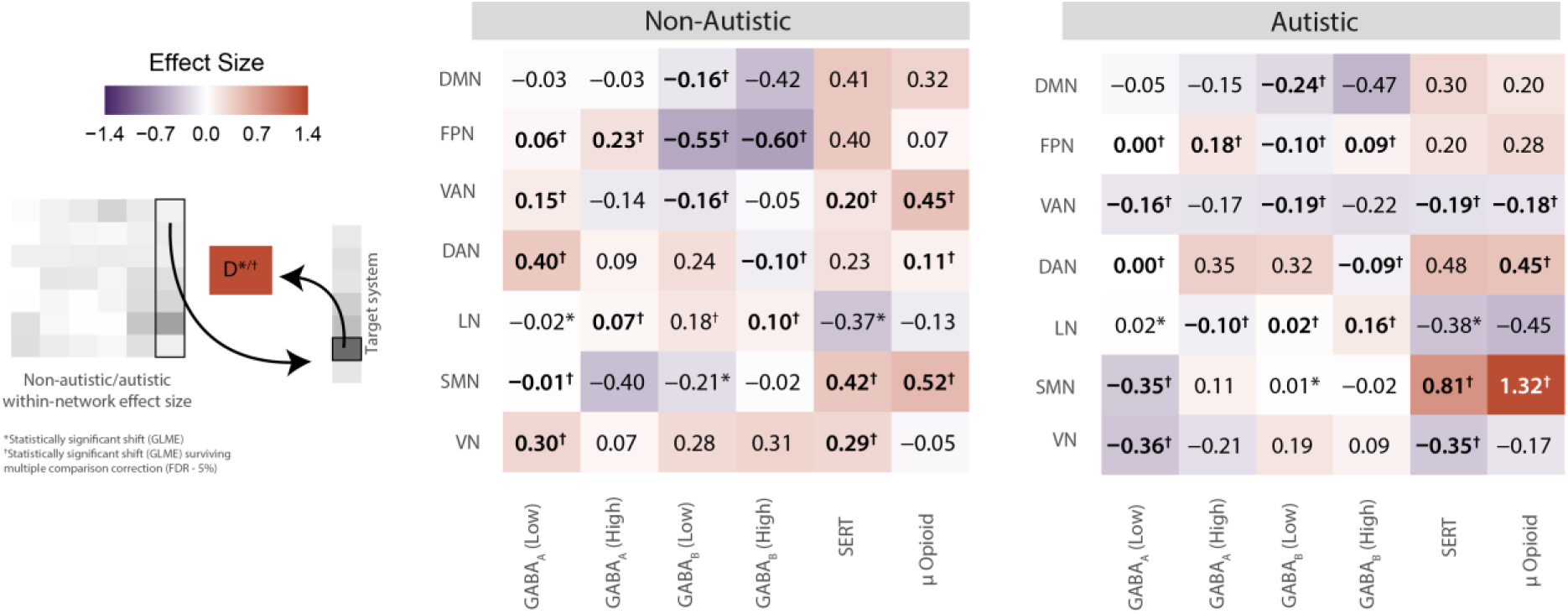
Within-network functional connectivity ‘shifts’ by neurotransmitter system targeted for the non-autistic and autistic groups. Values represent the effect sizes (Cohen’s d) and significant dose × group interactions are shown with an asterisk (uncorrected p < 0.05*, p obtained with a two-sided permutation test), those that survive FDR correction are shown in bold and with a dagger (p < 0.05^†^). DAN, dorsal attention; DMN, default mode; FPN, frontoparietal; LN, limbic; VAN, ventral attention & VN, visual network.

**Figure 3.**
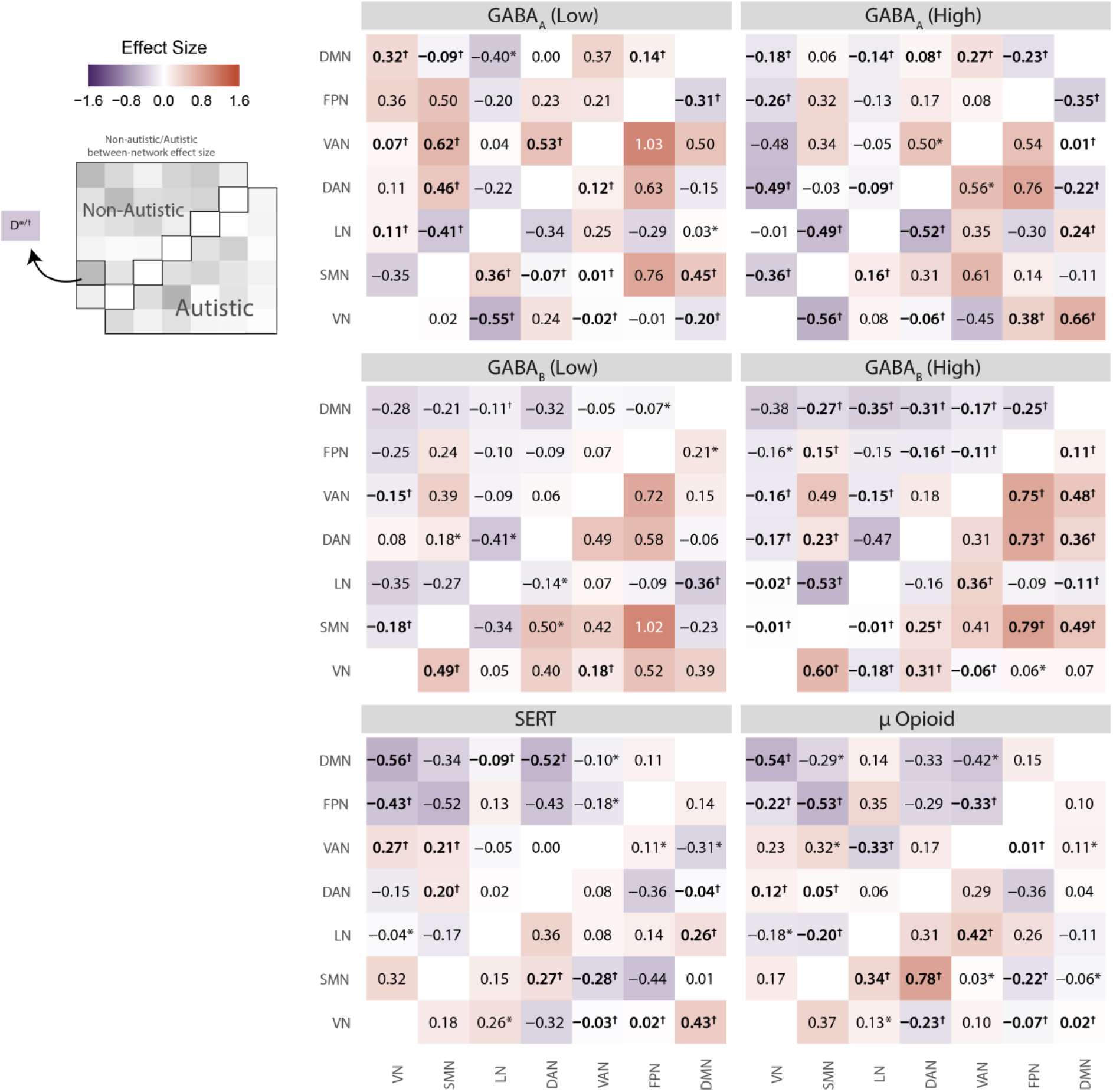
Between-network functional connectivity ‘shifts’ by neurotransmitter system targeted for the non-autistic and autistic groups. Values represent the effect sizes (Cohen’s d) and significant dose × group interactions are shown with an asterisk (uncorrected p < 0.05*, p obtained with a two-sided permutation test), those that survive FDR correction are shown in bold and with a dagger (p < 0.05^†^). DAN, dorsal attention; DMN, default mode; FPN, frontoparietal; LN, limbic; VAN, ventral attention & VN, visual network.

**Table 3.** Key dose × group interaction findings for within- and between-network connectivity across all drug challenges.

| Dose | Network | Non-Autistic Effect Size | Autistic Effect Size | P<sub>unc</sub> | P<sub>FDR</sub> |
| --- | --- | --- | --- | --- | --- |
| **GABA<sub>A</sub>** |  |  |  |  |  |
| *Within-networks* |  |  |  |  |  |
| 10 mg | VN | 0.30 | -0.36 | <0.001 | <0.001 |
| *Between-networks* |  |  |  |  |  |
| 10 mg | VN-DMN | 0.32 | -0.20 | <0.001 | <0.001 |
| 20 mg | VN-DMN | -0.18 | 0.66 | <0.001 | 0.001 |
| 20 mg | VN-FPN | -0.26 | 0.38 | <0.001 | <0.001 |
| **GABA<sub>B</sub>** |  |  |  |  |  |
| *Within-networks* |  |  |  |  |  |
| 15 mg | FPN | -0.55 | -0.10 | <0.001 | 0.003 |
| 30 mg | FPN | -0.60 | 0.09 | <0.001 | <0.001 |
| *Between-networks* |  |  |  |  |  |
| 30 mg | FPN-SMN | 0.15 | 0.79 | 0.015 | 0.025 |
| 30 mg | FPN-VAN | -0.11 | 0.75 | <0.001 | <0.001 |
| 30 mg | FPN-DAN | -0.16 | 0.73 | <0.001 | <0.001 |
| 30 mg | DMN-SMN | -0.27 | 0.49 | <0.001 | <0.001 |
| 30 mg | DMN-VAN | -0.17 | 0.48 | <0.001 | <0.001 |
| **Serotonin** |  |  |  |  |  |
| *Within-networks* |  |  |  |  |  |
| 20 mg | SMN | 0.42 | 0.81 | <0.001 | 0.002 |
| 20 mg | VN | 0.29 | -0.35 | <0.001 | <0.001 |
| 20 mg | VAN | 0.20 | -0.19 | 0.008 | 0.022 |
| *Between-networks* |  |  |  |  |  |
| 20 mg | DMN-VN | -0.56 | 0.43 | <0.001 | <0.001 |
| **μ-opioid** |  |  |  |  |  |
| *Within-networks* |  |  |  |  |  |
| 12.5 mg | SMN | 0.52 | 1.32 | <0.001 | <0.001 |
| 12.5 mg | VAN | 0.45 | -0.18 | <0.001 | <0.001 |
| *Between-networks* |  |  |  |  |  |
| 12.5 mg | LN-VAN | -0.33 | 0.42 | 0.002 | 0.005 |
| 12.5 mg | SMN-DAN | 0.05 | 0.78 | <0.001 | <0.001 |

#### Targeting the GABA system

##### Targeting GABA_A_

###### Within-network connectivity

Within-network connectivity was differentially shifted by GABA_A_ activation in each group across several networks. In particular, the low dose increased connectivity of the visual network in non-autistic participants but it decreased in autistic participants.

###### Between-network connectivity

In general, shifts in between-network connectivity elicited by GABA_A_ activation were more pronounced in the autism group compared to non-autistic participants.

In non-autistic participants, GABA_A_ activation by low and high dose AZD7325 elicited broadly similar shifts in between-network connectivity apart from between-network connectivity of the visual network, which was significantly reduced by the high dose only.

In autistic participants, GABA_A_ activation by low and high dose AZD7325 elicited a broadly similar pattern of shifts in between-network connectivity but again, the exceptions were mostly in between-network connectivity of the visual network, specifically, increases in connectivity with the higher-order default mode and frontoparietal networks at the higher dose.

Together, GABA_A_ activation primarily (but not exclusively) shifted within and between-network connectivity of the visual network. The most marked shifts in connectivity were present in the autistic group. Distinct decreases in between-network of higher-order networks (e.g. DMN) responses in the autism were evident at the low dose. By contrast, there were increases in connectivity between these networks at the high dose.

##### Targeting GABA_B_

###### Within-network connectivity

Within-network responses to GABA_B_ agonism were qualitatively similar in the autistic and non-autistic groups and at the low and high doses. Lower-order (visual) network connectivity tended to increase, and higher-order network connectivity tended to decrease in response to arbaclofen. However, within-network responses were more pronounced in the non-autistic group, particularly in the frontoparietal network.

###### Between-network connectivity

There was a differential response in between network connectivity in autistic and non-autistic groups across the whole brain. In non-autistics, GABA_B_ activation by low and high doses of arbaclofen predominantly decreased between-network connectivity. In autistics, low and high doses of arbaclofen predominantly increased between-network connectivity.

Thus, GABA_B_ activation had a more prominent impact within-networks in non-autistic compared to autistic participants. However, there was an opposite effect on between-network connectivity across the brain in autistic (increased connectivity) compared to non-autistic participants (decreased connectivity). These group differences were more marked at the higher dose and particularly pronounced in higher-order and attentional/sensory networks.

#### Targeting the serotonin system

##### Within-network connectivity

Within-network connectivity responses to serotonin activation were broadly similar in each group except for visual and ventral attention network connectivity which was increased in the non-autistic group and decreased in the autistic group.

##### Between-network connectivity

Activation of serotonin signalling primarily elicited a decrease in between-network connectivity of higher order networks (such as default mode and frontoparietal) in non-autistic but not and autistic participants; indeed, serotonin activation significantly increased between-network connectivity of the default mode network in autism and did the opposite in non-autistic participants.

#### Targeting the opioid system

##### Within-network connectivity

The direction of response to μ-opioid agonism within-networks was broadly similar in direction in both groups but of a greater magnitude in autistic participants. There was especially more prominent increase in somatomotor and decrease in limbic network connectivity in autistic participants compared to non-autistic participants. In contrast, ventral attention network connectivity was increased by μ-opioid activation in non-autistic participants groups but decreased in the autistic participants.

##### Between-network connectivity

Again, there were differential effects in each group on between-network connectivity. In general, μ-opioid activation increased between-network connectivity of somatomotor and limbic networks in the autism group. Whereas it decreased connectivity of the higher order default mode and frontoparietal networks in non-autistic participants.

Thus, activation of the μ-opioid system has similar effects on within-network connectivity and differential effects on between-network connectivity, with differences in autism concentrated upon lower-order (e.g. somatomotor), limbic and attentional within and between-network connectivity.

### The divergent ‘shift’ of the autistic brain in response to drug challenge

Next, we investigated whether there was an overarching pattern to the way in which the autistic brain responded to drug challenge overall (irrespective of the neurotransmitter system targeted) by pooling together the effect sizes for each pharmacological probe as reported above. Overall, the autistic brain had a greater magnitude of shift in response to any drug, compared to the non-autistic brain (median: non-autistic, −0.035 & autistic, 0.08; interquartile range: non-autistic, 0.40 & autistic, 0.48; p < 0.001). A net positive shift in connectivity was observed in autism (mean effect size 0.11), and a slight decrease in connectivity in non-autistic participants (mean effect size −0.03).

Post-hoc analyses confirmed that this group-level difference in network responsivity was primarily driven by *increases* in *between-network* connectivity in autism compared to a minimal decrease in the non-autistic group (median: non-autistic, −0.09 & autistic, 0.11; interquartile range: non-autistic, 0.41 & autistic, 0.45; p = <0.0001); and there was no difference in within-network connectivity shifts between groups (median: non-autistic, 0.07 & autistic, −0.01; interquartile range: non-autistic, 0.39 & autistic, 0.38; p = 0.20). Ventral attention, frontoparietal, somatomotor and (to a lesser extent) dorsal attention network connectivity increases are the largest contributors to shifts in between-network connectivity in autism; whereas, default mode, limbic and visual network connectivity remained unchanged (or were slightly increased) (**Fig. 4**).

**Figure 4.**
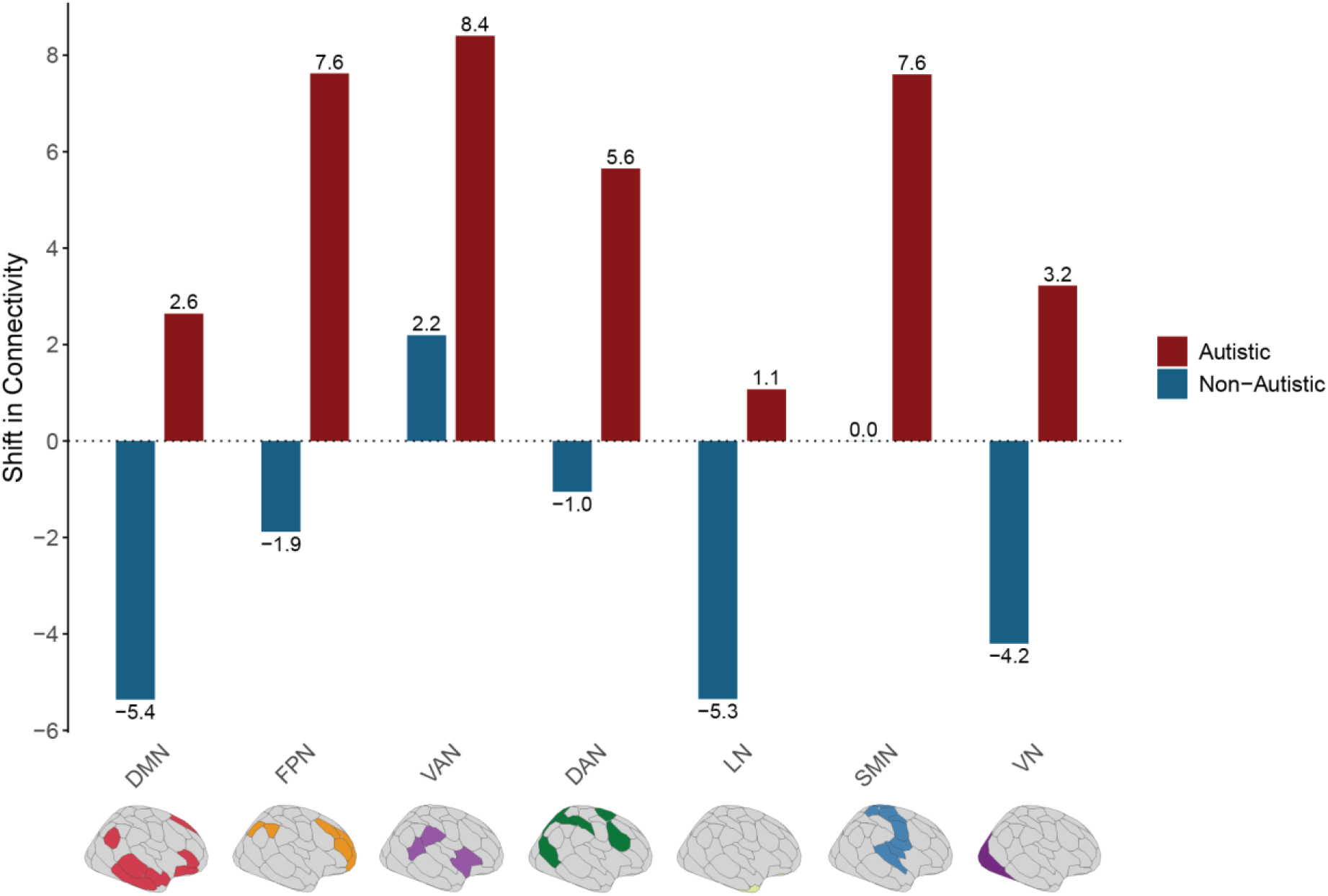
Aggregate between-network connectivity shifts (values represent Cohen’s d effect size) for each resting-state network separately for both groups. A distinct pattern of increased connectivity in autism is observed, and largely reduced connectivity in non-autistic participants across networks in response to drug challenge. DAN, dorsal attention; DMN, default mode; FPN, frontoparietal; LN, limbic; VAN, ventral attention & VN, visual network.

## Discussion

We have used a uniform analytical framework to capture the change in functional connectivity within and between seven brain networks following activation of several known targets of the GABA_A_, GABA_B_, serotonin and μ-opioid neurotransmitter systems in autistic and non-autistic adults. As predicted, despite minimal baseline group differences, functional connectivity measures were ‘shifted’ differently by each drug probe in autistic compared to non-autistic individuals.

There were some target-specific patterns; modulation of GABA_A_ shifted visual network connectivity; GABA_B_ shifted between-network connectivity across the brain; serotonin shifted sensory and higher order networks (e.g. default mode network); and μ-opioid shifted somatomotor and limbic network connectivity. However, we also found that, regardless of neurotransmitter system targeted, in general drugs elicited significantly greater increases in between-network connectivity in autistic participants compared to their non-autistic peers; especially between-network connectivity of the VAN, FPN and SMN, which are primarily networks which ‘allow’ bottom-up information to shift attention and control the initiation of behaviour^31,32^.

### Target-specific patterns of connectivity shift are different in autistic and non-autistic participants

We observed target-specific differences in functional connectivity responses in autism depending on the neurotransmitter system probed. The networks which shifted in response to each drug broadly aligned with the known expression pattern of their receptors but the direction and/or extent of shift in connections within and between these brain regions was different in autism.

#### GABA response differences

Autistic differences in network responsivity following GABA_A_ activation were observed in lower-order sensory networks. The effect was most prominent in the visual cortex where GABA_A_ receptor expression is reported to be particularly high^33^. Altered responsivity in autism may, in part, be explained by reduced levels of GABA_A_ receptors in the autistic brain^34,35^.

However, this evidence comes from post-mortem data, and the reported decrease in GABA receptors has been less consistent in the living human brain using PET imaging. Recent PET data suggests that cortical GABA_A_ receptor levels may be no different in autistic and non-autistic adults^36,37^.

There were more widespread autistic differences in between-network connectivity in response to GABA_B_ activation, and this may reflect the broader extent of GABA_B_ expression across the whole brain^33,38^. Again, this may, in part, be explained by reduced levels of GABA_B_ receptors in the autistic brain^34,35^. For example, GABA_B_ receptors may be differently expressed in prefrontal and parietal regions that comprise higher-order and attentional networks in which we see the largest differential shifts in response to arbaclofen here^39^. However, there are no reports to-date of altered GABA_B_ receptor expression in autism from PET data. Thus, minimal baseline differences in GABA receptor availability may not entirely explain the diverging direction of shifts in connectivity of autistic and non-autistic resting-state networks in response to GABA_A/B_ activation.

We suggest that the explanation may be functional differences in the GABA system. This is consistent with other evidence from preclinical animal work and humans. For example, agonism of GABA_A_ and GABA_B_ receptors ameliorates autistic features in an autism-relevant mouse model^40^; and we have shown that visual and auditory sensory processing in the same autistic and non-autistic adults who participated in the present study are also differentially modulated by GABA_B_ receptor agonism^15,41^.

#### Serotonin response differences

There were paradoxical responses to citalopram in the autistic brain. In contrast to non-autistic participants, within-network connectivity of the visual and ventral attention networks was reduced in the autistic brain, but connectivity between the visual and default mode network was increased.

In the visual network, there is evidence for lower SERT levels in the visual cortex in autism^42^, several regions of the DMN also express SERT including the medial prefrontal cortex and posterior cingulate cortex (PCC)^43^ and lower SERT binding has been reported in these regions in autistic adults^44^. However, it is difficult to align receptor availability differences with the range of within and between-network alterations elicited by citalopram in autism. The increase in between-network connectivity of the DMN in autism in response to citalopram is noteworthy given that altered DMN connectivity is implicated in depression, which is more common in autistic people^45^. The augmented responses to SERT activation in autistic individuals across the default mode and sensory (e.g. visual) networks observed here aligns with evidence that autistic patients respond at lower doses to SSRIs^45^ and SSRIs can be poorly tolerated by autistic people^46^. We do however acknowledge that this is a single dose in which citalopram was used as a pharmacological probe to elevate bulk serotonin levels to understand the role of the serotonin system and not intended to directly extend to the mechanistic underpinnings of anti-depressant efficacy in autistic and non-autistic people – where treatment with citalopram often requires longer term use (6-8 weeks) and may therefore have different network-level effects than those reported here.

#### μ-opioid response differences

Lastly, in autistic participants, agonism at the μ-opioid receptor by tianeptine altered within and between-network differently in autistics. Within-network connectivity was increased in somatomotor networks and decreased in limbic and ventral attention networks in autistics but not in non-autistics. Between-network connectivity of the ventral attention and limbic network was increased in autism and decreased in non-autistic participants.

μ-opioid receptors are expressed in several components of these networks with particularly high expression in the cingulate cortex of the limbic network^47^ and in somatomotor regions^48^. Recent human PET data suggests that baseline differences in cortical μ-opioid receptor expression in autism are restricted to upregulation in the precuneus only^49^, so again differences in receptor levels do not explain responsivity differences. Thus, as for other drug probes, what sets the autistic group apart is a distinct functional response of the brain to neurotransmitter perturbation.

These observed differences in the responsivity of resting-state networks across the neurotransmitter systems probed here may need to be understood in the context of neurodevelopment. Indeed, we have evidence for early alterations in functional connectivity in newborn infants who have a higher likelihood of a neurodevelopmental outcome such as autism^50^. This work observed that local functional connectivity in neonates with a higher likelihood of autism was greater in sensory cortices and limbic areas responsible for face processing^51^. We have more recently reported that moment-to-moment dynamic functional connectivity profile of brain states that engage sensorimotor networks at birth predicts neurodevelopmental and autistic traits at 18 months^52^. Thus, by birth, there are already differences in the functional landscape of brain regions associated with autism. Although, capturing convincing baseline cross-sectional differences in functional connectivity in larger autistic and non-autistic cohorts assessed at later ages has been challenging^3^. We suggest that what persists in autism is not any average baseline difference in functional connectivity, but a true functional difference as defined by altered response to perturbation (in this case by pharmacological probe).

Consistent with this, we see no substantial baseline difference in resting-state network connectivity between groups of adults in our sample. It could be argued that this is unsurprising given our relatively small sample size but one recent mega-analysis used a sample of 1824 individuals, 796 of whom were autistic to report effects which were still relatively small^7^. What is more striking therefore, is that significant differences in functional connectivity responses to drug challenge were observed even with our small sample size.

#### Between network functional connectivity in autism increases in response to drug challenge

We pooled resting-state connectivity data across all the pharmacological probes to investigate if there was any generalised response profile common to the autistic group. We report that overall, drug challenge *increased between-network connectivity in autism*, but either had little impact or *decreased* between-network connectivity in non-autistic participants. The main drug-elicited increases in functional connectivity were mainly localised to between-network connectivity of the VAN, FPN and SMN, which are implicated in detecting unexpected stimuli to trigger attentional shift, flexible initiation of new tasks and the processes controlling motor output^53^, respectively. Thus, to some extent they can be thought of as allowing a flexible response to external stimuli. Consequently, how neurotransmitter systems in autistic individuals regulate the engagement and disengagement of functional networks in response to changing task demands may be altered. By facilitating connectivity between-networks the drugs used here may promote more flexible responses to external stimuli. We emphasise however that this is highly speculative and whether this profile has clinical meaning remains to be examined.

An additional or alternative explanation for the increased between-network shift induced by drug challenge in autistic participants may be that homeostasis is differently regulated in autism. For example, if networks remain in their shifted state for longer after drug challenge in autistic participants, we may simply be continuing to capture a functional shift in autistics which has moved back towards baseline in non-autistic participants by the time of data acquisition. In support of this, we have previously reported that limbic system activation takes longer to return to baseline in autism following citalopram during a fMRI face processing task^54^. This result was restricted to *between*-networks, so following perturbation local systems may return to baseline quicker compared to those *between*-networks that are more distal and/or more functionally segregated. Our previous work using other modalities to capture brain dynamics supports this position. We have used the aperiodic (1/f) slope of EEG to show that, despite no baseline group differences, autistic participants ‘shifted’ their 1/f slope at lower doses than non-autistic participants^55^. Placed together with the present analyses, the homeostatic mechanisms which might be at play in non-autistic people to limit shifts in brain function are not so evident in autism. MRI does not allow us to address this question but there is some evidence from cellular models that synaptic scaling to adjust excitatory or inhibitory synaptic strength up or down to stabilise neural firing rates^56^ is disrupted in animal models of autism^57^.

We acknowledge several limitations to our work. First, in humans we are often limited to non-selective pharmacological probes (i.e. with more than one molecular target). While both GABA probes used here were relatively selective compounds. For example, tianeptine also has agonist activity at delta (δ) opioid receptors^58^ and indirect effects on other neurotransmitter systems (e.g. glutamate^59^ and serotonin^60^) which may also contribute to its impact on functional connectivity. Second, our approach included different MRI acquisition sequences adopted between studies (SERT/ μ-opioid versus the GABA probes). The impact of this was somewhat mitigated by using a consistent preprocessing pipeline across all studies. Similarly, an atlas-based parcellation approach was used to provide a uniform analysis framework across studies/probes. Third, motion artifacts (i.e. due to participants moving in the scanner) can alter functional connectivity measures^61^. As a result, our preprocessing pipeline included several techniques focused on motion correction specifically (e.g. despiking, multi-echo ICA correction, censoring). There was also stringent post-processing quality control and datasets with significant movement (or other quality control issues) were omitted from analyses. Fourth, our sample size for each pharmacological probe is relatively small and the sexes are also not balanced (only males were recruited for the studies targeting SERT and the μ-opioid receptor), therefore more work including female needs to be done. This is a recognised problem in past pharmacological research^62^ and in our current studies our goal is to recruit an equal number of men and women. Despite a small sample size for each probe our statistical power was increased due to the within-subject repeated-measures design of our studies.

In conclusion, our findings suggest that the modulation of resting-state network connectivity by multiple key neurotransmitter systems is different in autistic and non-autistic participants. These systems are targeted by medications used to treat neurological and psychiatric conditions experienced by neurodivergent people. Our work indicates that the autistic brain is ‘pharmacologically distinct’. This is crucial because autistic people have a higher incidence of psychiatric and neurological conditions^63,64^ but drug development and trials for mental health problems do not consider neurodivergence. These results suggest this needs to change. Investigating if these drug-induced ‘shifts’ in functional connectivity can help provide better targeted interventions will be an important next translational step.

## Data Availability

Data from this study is available upon request.

## Acknowledgements

Competing Interests

T.P.W. holds shares in Compass Pathfinder Ltd. N.A.P. has consulted for Deerfield Discovery and Research. D.G.M.M. has consulted for Jaguar Gene Therapy LLC. G.M.M. has received funding for investigator-initiated studies from GW Pharmaceuticals and Compass Pathfinder Ltd. G.M.M. has consulted for Greenwich Biosciences, Inc. L.G.S.F., M.D., C.M.P., H.V., A.C.P., C.L.E., G.I., J.L.F., R.H.W., F.M.P., J.K., N.M., N.M.L.W., D.B. and E.D. have no conflicts to declare.

## Author Contributions

T.P.W. performed data preprocessing and analysis, prepared and drafted the manuscript. L.G.S.F. contributed to data analysis and helped draft the manuscript. M.D. contributed to data collection and preprocessing, and helped draft the manuscript. C.M.P., H.V., A.C.P., C.L.E., J.L.F., R.H.W., F.M.P., J.K., N.M.L.W. contributed to data collection. G.I. provided pharmacy services for the studies. N.M. contributed to preprocessing and data analysis. D.B. and N.A.P. edited the manuscript and provided input on data analysis. E.D., D.G.M.M. and G.M.M. conceived the idea for the studies; contributed to study design and drafted the manuscript. All authors reviewed and approved the final manuscript.

## Data and Material Availability

Data from this study is available upon request.

## Supplementary Information

**Supplementary Table 1.** Demographic information of the sample. Values in brackets represent the standard deviation.

| GABA: AZD Study |  |  |  |  |
| --- | --- | --- | --- | --- |
|  | Non-Autistic | Autistic | statistic | p value |
| Age | 35 (± 13) | 34 (± 10) | t = 0.12 | p = 0.91 |
| IQ | 120 (± 13) | 112 (± 12) | t = 1.53 | p = 0.14 |
| GABA: Arbaclofen Study |  |  |  |  |
|  | Non-Autistic | Autistic | statistic | p value |
| Age | 28 (± 8) | 34 (± 10) | -2.64 | p = 0.01 |
| IQ | 120 (± 10) | 117 (± 10) | 1.33 | p = 0.19 |
| Serotonin & Opioid Study |  |  |  |  |
|  | Non-Autistic | Autistic | statistic | p value |
| Age | 26 (± 7) | 29 (± 10) | w = 150.5 | p = 0.39 |
| IQ | 114 (± 10) | 111 (± 16) | t = 0.66 | p = 0.52 |

**Supplementary Table 2.** Additional MRI acquisition information.

| Study ID | Scan | Inversion Time (ms) | Flip Angle (°) | Field of View (mm) | Matrix Size (no. of voxels) | No. of Sagittal Slices | Axial Slices per Volume | Slice Thickness (mm) | Interslice Gap (mm) | Voxel size | No. of volumes |
| --- | --- | --- | --- | --- | --- | --- | --- | --- | --- | --- | --- |
| GABA | Structural | 400 | 11 | 270 | 256 x 256 | 196 | - | 1.2 | 0 | 1.05 x 1.05 x 1.2 mm | 1 |
|  | Functional | 0 | 80 | 240 | 64 x 64 | - | 32 | 3.0 | 1.0 | 3.75 x 3.75 x 3.0 mm | 192 (length = 8 mins) |
| Serotonin & Opioid | Structural | 400 | 11 | 270 | 256 x 256 | 196 | - | 1.2 | 0 | 1.055 x 1.055 x 1.2 mm | 1 |
|  | Functional | 0 | 90 | 240 | 64 x 64 | - | 33 | 3.8 | 0.4 | 3.75 x 3.75 x 4.5 mm | 215 (length = 8.24 mins) |

**Supplementary Table 3.** Degree of head motion in the non-autistic and autistic groups across all studies. Mean FD, mean framewise displacement.

| Study | Condition | Dosage (mg) | Non-Autistic Mean FD (mm) | Autistic Mean FD (mm) | Test Statistic | P Value |
| --- | --- | --- | --- | --- | --- | --- |
| GABA | Placebo | 0 | 0.07 (± 0.03) | 0.07 (± 0.04) | 59 | 1.00 |
| | AZD | 10 | 0.07 (± 0.05) | 0.10 (± 0.10) | 57 | 0.642 |
| | AZD | 20 | 0.06 (± 0.03) | 0.13 (± 0.14) | 62 | 0.583 |
| | Placebo | 0 | 0.15 (± 0.15) | 0.18 (± 0.09) | 105 | 0.117 |
| | Arbaclofen | 15 | 0.15 (± 0.12) | 0.22 (± 0.16) | 121 | 0.156 |
| | Arbaclofen | 30 | 0.16 (± 0.15) | 0.19 (± 0.15) | 212 | 0.326 |
| Serotonin & opioid | Placebo | 0 | 0.10 (± 0.05) | 0.15 (± 0.08) | 97 | 0.026 |
| | Citalopram | 20 | 0.12 (± 0.06) | 0.12 (± 0.06) | 49 | 0.528 |
| | Tianeptine | 12.5 | 0.11 (± 0.04) | 0.13 (± 0.05) | 110 | 0.106 |

**Supplementary Table 4.** The main effect of group showing differences in connectivity of all resting-state networks between autistic and non-autistic participants at baseline (placebo). P values were corrected for multiple comparisons across all networks.

| Network | | $P_{unc}$ | $P_{FDR}$ |
| --- | --- | --- | --- |
| SMN | VN | 0.0066 | 0.1848 |
| SMN | SMN | 0.054 | 0.54145 |
| LN | SMN | 0.0717 | 0.54145 |
| VAN | VN | 0.0807 | 0.54145 |
| VAN | VAN | 0.0986 | 0.54145 |
| DAN | DAN | 0.1233 | 0.54145 |
| SMN | VAN | 0.1492 | 0.54145 |
| DMN | LN | 0.1547 | 0.54145 |
| LN | LN | 0.2466 | 0.6776 |
| LN | VAN | 0.2751 | 0.6776 |
| DAN | LN | 0.2929 | 0.6776 |
| DAN | SMN | 0.3099 | 0.6776 |
| FPN | LN | 0.3146 | 0.6776 |
| DMN | FPN | 0.3843 | 0.726075 |
| DMN | DMN | 0.4072 | 0.726075 |
| FPN | FPN | 0.4149 | 0.726075 |
| DAN | VN | 0.4613 | 0.75978 |
| FPN | VAN | 0.5147 | 0.80064 |
| FPN | VN | 0.5708 | 0.841179 |
| DMN | SMN | 0.6525 | 0.9135 |
| DAN | VAN | 0.708 | 0.923364 |
| FPN | SMN | 0.7255 | 0.923364 |
| DAN | DMN | 0.7674 | 0.934226 |
| VN | VN | 0.8265 | 0.950432 |
| LN | VN | 0.8486 | 0.950432 |
| DAN | FPN | 0.9452 | 0.9738 |
| DMN | VN | 0.9518 | 0.9738 |
| DMN | VAN | 0.9738 | 0.9738 |

**Supplementary Figure 1.**
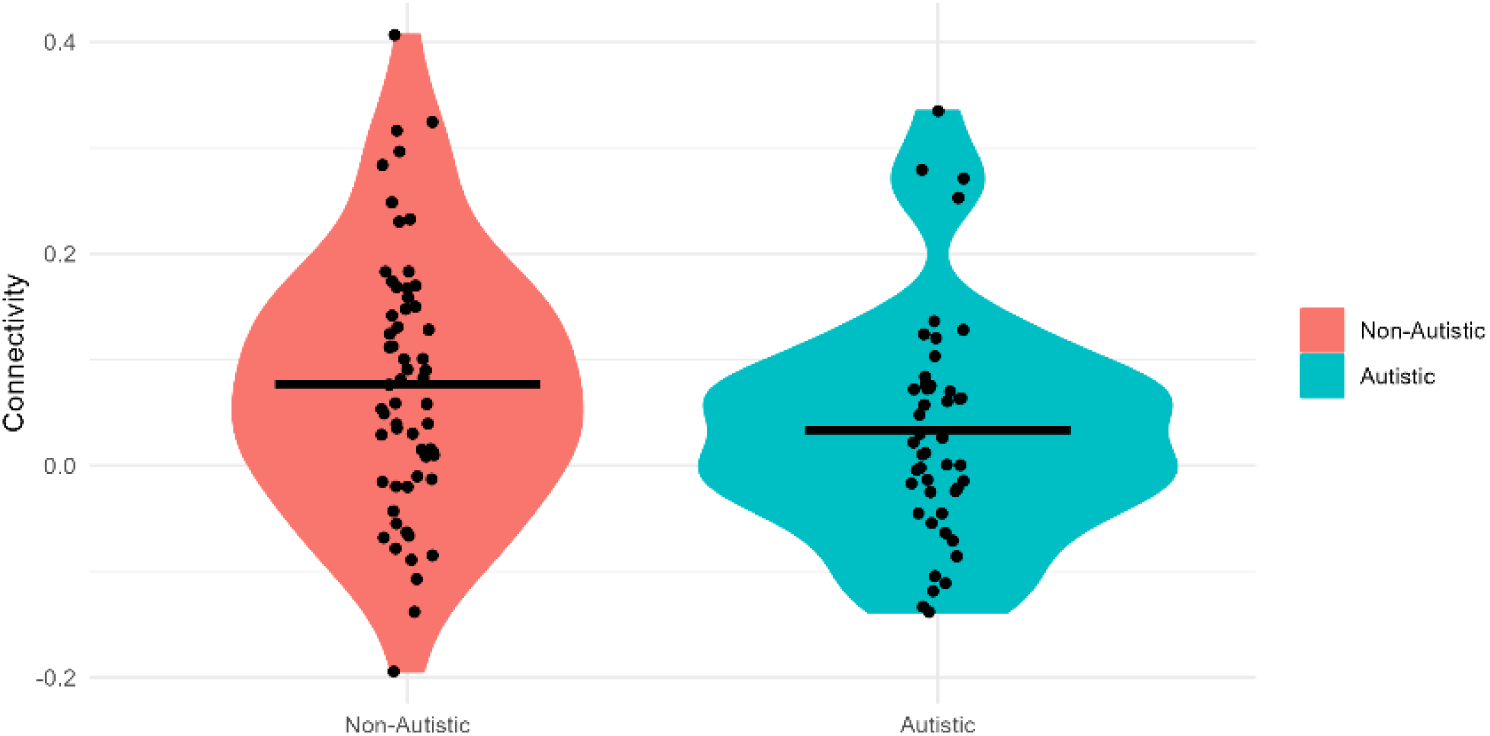
Baseline difference in somatomotor-visual network connectivity between the non-autistic and autistic groups.

